# Preoperative Quality-of-Life and Recovery Following Esophagectomy: Identifying Modifiable Factors Associated with Improvement

**DOI:** 10.1101/2025.07.30.25332378

**Authors:** Samir Amin, Joshua Cheruvathur, Jin Kweon, Mehrnoush Dehghani, Sara Najmeh, Carmen Mueller, Jonathan Cools-Lartigue, Lorenzo Ferri, R. Trafford Crump

## Abstract

**BACKGROUND:** Esophagectomy remains the primary curative treatment for esophageal cancer, but it often leads to substantial declines in health-related quality of life (HRQoL). While prior studies have identified various demographic and clinical factors associated with postoperative HRQoL, most are non-modifiable and offer limited opportunities for intervention. This study aimed to examine whether modifiable preoperative factors, particularly baseline HRQoL, are associated with clinically meaningful improvement in HRQoL one year after esophagectomy.

**METHODS:** This retrospective cohort study used data from the McGill Esophageal and Gastric Database. Patients were included if they underwent curative esophagectomy for esophageal cancer and completed the Functional Assessment of Cancer Therapy-Esophageal (FACT-E) both before surgery and at one year postoperatively. HRQoL improvement was defined as a ≥ 5-point increase in total FACT-E score. Stepwise multivariable logistic regression was used to evaluate the association between preoperative factors and HRQoL improvement, with additional sensitivity analyses across various improvement thresholds.

**RESULTS:** Of 108 eligible patients, 42% experienced clinically meaningful HRQoL improvement one year after surgery. Lower preoperative FACT-E scores were significantly associated with greater odds of improvement. No other modifiable factors, including BMI, smoking, or alcohol use, were significantly associated with improvement. Sensitivity analyses confirmed consistent findings across alternative HRQoL thresholds. Among demographic variables, female sex was associated with lower odds of improvement.

**CONCLUSION:** Patients with greater preoperative symptom burden were more likely to experience meaningful improvements in HRQoL after esophagectomy. These findings support the value of routine preoperative HRQoL assessment and suggest that baseline HRQoL could be used to identify patients who may benefit from targeted prehabilitation or psychosocial support. As precision medicine advances, incorporating patient-reported outcomes into preoperative risk stratification could enhance personalized recovery planning and improve survivorship care.

## INTRODUCTION

Esophagectomy remains the primary curative treatment for esophageal cancer, but it is a complicated operation associated with significant morbidity (1). Many patients experience serious postoperative complications that affect their ability to eat, swallow, and carry out daily activities (2). As a result, esophagectomy often has a profound effect on patients’ overall health-related quality of life (HRQoL) (2).

In recent years, HRQoL has emerged as a key outcome in cancer research, complementing traditional endpoints like survival (3). HRQoL captures a patient’s overall well-being across physical, emotional, social, and functional domains, as shaped by both disease and treatment (3). It is most often measured through patient-reported outcome measures (PROMs), which allow patients to directly report on their symptoms, daily functioning, and overall health status (4). In the context of esophageal cancer, where treatment can be physically and emotionally demanding, some have argued that HRQoL may be as important as survival—if not more so—in guiding care decisions and evaluating outcomes (2).

The trajectory of HRQoL following esophagectomy is known to be complex and variable. Studies show that patients experience an initial decline in HRQoL during the early postoperative period, as measured by PROMs, due to surgical recovery and treatment side effects (5). Over time, however, many survivors improve. By one year post surgery, HRQoL often recovers to baseline levels or even exceeds pre-treatment scores for a substantial proportion of patients (5). This recovery aligns with the clinical impression that successful removal of the tumour (and relief of dysphagia or pain) eventually improves patients’ well-being.

However, not all patients follow this optimistic trajectory. Some studies report persistent HRQoL deficits in certain survivors, with one analysis noting that average HRQoL never fully returned to baseline even nine years post-esophagectomy (6). Such findings underscore that there is significant heterogeneity in patients’ HRQoL: while some individuals rebound after surgery, others continue to struggle with symptoms like fatigue, eating difficulties, or reduced physical and social function. This variability in outcomes has made postoperative HRQoL an important clinical concern, highlighting the need to understand why some patients fare better than others. By identifying what factors predispose patients to better or worse HRQoL after esophagectomy, clinicians hope to improve preoperative counselling, manage expectations, and tailor interventions to support recovery.

Several studies have examined factors associated with HRQoL following esophagectomy (7) (8) (9). Crucially, however, nearly all the factors identified are unmodifiable (i.e., factors that cannot be altered through medical intervention or lifestyle changes). For example, a patient’s age, sex, or tumor stage cannot be changed prior to surgery. While knowing these factors can help assess risk, they offer limited practical avenues to improve outcomes. In other words, identifying a high-risk patient is not the same as being able to alter that patient’s risk profile. This limitation has been noted as a challenge in HRQoL research; without actionable associations, HRQoL data provide insight but little direct benefit at the bedside (10).

The purpose of this study is to identify preoperative modifiable factors that are associated with post-esophagectomy HRQoL improvement. In particular, we sought to identify which modifiable patient or clinical characteristics present before surgery are associated with an improvement in HRQoL one year after esophagectomy.

This study aims to address a key gap in the literature by identifying preoperative, modifiable factors associated with HRQoL improvement one year after esophagectomy. The results from this study could be a first step toward helping clinicians better identify patients who may benefit from targeted interventions, such as prehabilitation, counseling, or other lifestyle changes that maximize their post-esophagectomy HRQoL outcomes.

## METHODS

### Study Design

We conducted a retrospective cohort study using routinely collected clinical and patient-reported data from the McGill Esophageal and Gastric Data-Biobank (EGDB), a prospectively maintained clinical database that includes esophageal cancer patients treated between 2006 and 2025 by the Division of Thoracic and Upper Gastrointestinal Surgery at McGill University in Monreal, Canada (i.e., the Division). The Division is a high-volume tertiary care referral center for esophageal cancer in Quebec, Canada, serving a publicly insured population of approximately 8 million residents.

All patients included in this study provided consent to have their data used in secondary research. This study’s protocol was approved by the McGill University Health Centres Research (MUHC) Ethics Board (#2025-10674).

### Participants

Patients were eligible for inclusion if they were 18 years of age or older, had a confirmed diagnosis of esophageal cancer, underwent esophagectomy with curative intent, and initiated the Functional Assessment of Cancer Therapy–Esophageal (FACT-E) questionnaire both pre-esophagectomy and at one-year post-esophagectomy. Patients who initiated but did not complete either the pre- or one-year post-operative FACT-E with a scorable total score (i.e., insufficient responses for valid scoring) were excluded from the analysis.

### Study Size

As this was a hypothesis generating study, no a priori sample size calculation was performed. Instead, the study sample size was determined by the number of patients in the EGDB who met all eligibility criteria.

### Variables

The primary outcome of interest was improvement in HRQoL, defined as a ≥ 5-point increase in total FACT-E score between the pre-esophagectomy and the one-year post-esophagectomy time points. This threshold corresponds to the minimally important difference established in prior literature (11).

The FACT-E is a 44-item questionnaire used to assess HRQoL in patients with esophageal cancer (12). It includes four general cancer domains: physical (PWB), social/family (SWB), emotional (EWB), and functional well-being (FWB), plus a disease-specific subscale for esophageal cancer symptoms (ECS) (12). Each item is rated on a 5-point Likert scale based on symptom severity or impact over the past 7 days (12).

Scores are calculated using a standardized scoring algorithm with higher scores representing better HRQoL (12).The total FACT-E score ranges from 0 to 176, with domain-specific ranges of 0–28 for physical, social, and functional well-being, 0–24 for emotional well-being, and 0–68 for the esophageal cancer subscale (12).

The FACT-E was administered in either French or English (based on participant preference) at the initial consultation and again closer to the time of surgery, as well as at multiple post-operative timepoints, including one year after esophagectomy. For this study, the pre-esophagectomy FACT-E score was defined as the assessment completed at initial consultation when available. If unavailable, the closest available FACT-E assessment prior to surgery was used instead.

Pre-esophagectomy modifiable factors assessed for their association with HRQoL improvement included the pre-esophagectomy total FACT-E score and its five subscales: PWB, SWB, EWB, FWB, and the ECS. Additional modifiable factors included body mass index (BMI), smoking status, and alcohol consumption. These factors were selected based on prior evidence linking them to surgical outcomes and their potential for intervention through prehabilitation or behavioral support (2)(13)(14).

Potential confounders considered in the analysis included sex, age, Charlson Comorbidity Index (CCI), surgery type, pathological stage, tumor level, histologic subtype, and neoadjuvant treatment type. These were included to adjust for clinical and demographic differences known to influence postoperative recovery and quality of life outcomes (15)(16)(7)(8)(17).

### Data Sources and Measurement

All data were entered into a secure REDCap database hosted by the Research Institute of the MUHC. A de-identified dataset was provided to the research team for analysis. The EGDB includes data pertaining to participants’ demographic, clinical, pathological, and treatment-related information. These data were collected either directly from participants or abstracted from hospital medical records by trained data entry staff.

Efforts were made to minimise potential sources of bias throughout data collection and measurement. Measurement bias was reduced by using the validated FACT-E questionnaire, administered at consistent timepoints in the participant’s preferred language. Lifestyle and clinical variables were abstracted from electronic medical records by trained personnel using standardised procedures to ensure consistency across participants. Selection bias was minimized by the high enrolment rate into the EGDB among newly referred esophageal cancer patients at our center. However, patients with missing HRQoL data were excluded from the analysis, which may introduce bias if the missingness was not random.

### Statistical Methods

Descriptive statistics were used to summarise participants’ tumor and treatment characteristics, as well as pre-esophagectomy and 1-year post-esophagectomy HRQoL scores. Continuous variables were reported using means and standard deviations. Categorical variables were summarised using counts and percentages.

The primary outcomes, improvement in HRQoL, was categorised as a binary variable (0 = if FACT-E total score < 5 points, 1 = otherwise). All quantitative variables were analysed on their original scales unless otherwise specified. Pre-operative total FACT-E scores and subscale scores (PWB, SWB, EWB, FWB, and ECS) were scored according to the FACT-E guidelines and treated as continuous variables (12). BMI was treated as a continuous variable to preserve statistical power. CCI was treated as a categorical variable with four levels: none (0), mild (1–2), moderate (3–4), and severe (≥5). Age was modeled as a continuous variable. No transformations were applied to any variables. A linear relationship was assumed for all continuous variables.

The primary analysis assessed the association between modifiable pre-esophagectomy factors and improvement in HRQoL at 1-year post-esophagectomy. Associations were first examined using univariate logistic regression. A multivariable logistic regression model was then constructed in a stepwise fashion: beginning with modifiable factors, followed by tumor characteristics (e.g., level, histology, pathological stage), treatment-related variables (e.g., neoadjuvant treatment type, surgery type), and finally patient demographics (e.g., age, sex, Charlson Comorbidity Index). This modelling strategy was chosen to evaluate the independent contribution of modifiable factors while sequentially adjusting for potential clinical confounders.

To evaluate and compare model performance across the four stepwise multivariable logistic regression models, we calculated the Akaike Information Criterion (AIC), area under the receiver operating characteristic curve (AUC), and pseudo R² for each model. AIC was used to assess model fit while penalizing for complexity, with lower values indicating better fit (18). AUC provided a measure of discriminative ability, where values closer to 1.0 indicate better model performance and values around 0.5 suggest no better than chance (19). Pseudo R² offered an estimate of explained variance, with higher values reflecting greater explanatory power, though these values are typically lower than those observed in linear regression models (20).

Multicollinearity between variables was assessed using variance inflation factors (VIFs). Missing data were handled using median imputation for continuous variables and multiple imputation by chained equations (MICE) for categorical variables, allowing inclusion of participants with partially missing data. Sensitivity analyses were conducted to evaluate the robustness of results across varying thresholds for defining HRQoL improvement, using a range spanning the minimally important difference (4–9 points on the FACT-E scale) (11).

All statistical analyses were conducted using Python. A two-sided p-value of <0.05 was considered statistically significant. The code for this analysis is publicly available (https://github.com/Crump-Lab/ESO_project/blob/main/notebooks/ESO_ED_analysis_pre_n_baseline_n_1yrpost_4.ipynb).

## RESULTS

### Sample

The initial dataset extracted from the EGDB included 1,773 participants. Of these, 151 participants were excluded because they underwent endoscopic procedures (endoscopic submucosal dissection or endoscopic mucosal resection) rather than esophagectomy. An additional 101 participants were excluded because their surgeries were performed with palliative rather than curative intent. A further 1,397 participants were excluded because they did not initiate a pre-esophagectomy and one-year post-esophagectomy FACT-E. Finally, 16 patients were excluded for not completing both the pre- and post-esophagectomy questionnaires beyond the allowable threshold for missing data. The ultimate sample was comprised of 108 participants. Figure 1 illustrates how the sample was calculated.

**Figure 1.**
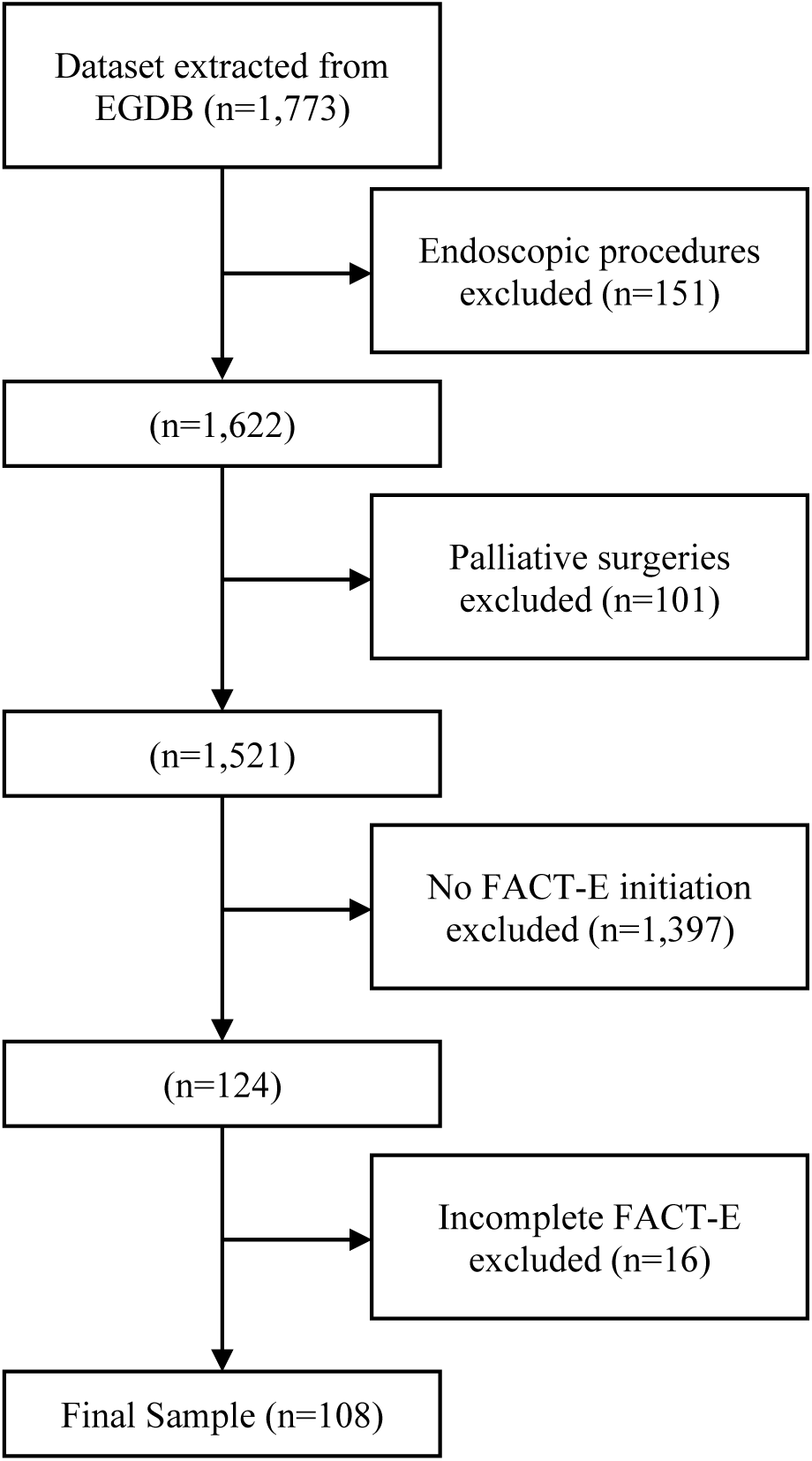
Flowchart of Participant Inclusion and Exclusion Criteria for Final Analytic Sample.

Demographic, clinical, and treatment characteristics of the sample can be found in Table 1. Of the 108 participants, 45 (41.7%) experienced a clinically significant HRQoL improvement at one year. There were no notable differences between improvers and non-improvers in demographics, lifestyle factors, tumor characteristics, or treatment exposures.

**Table 1.**
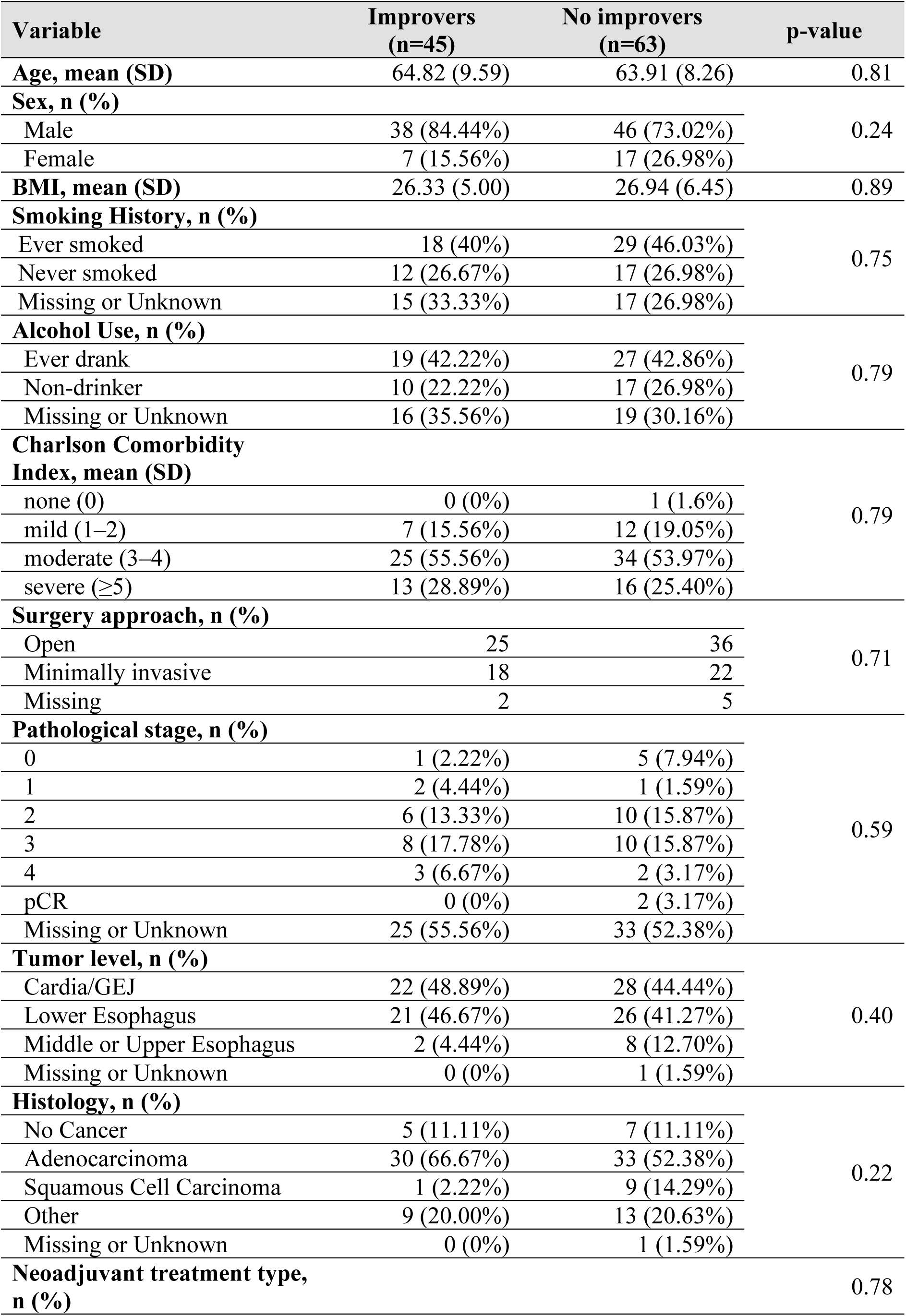
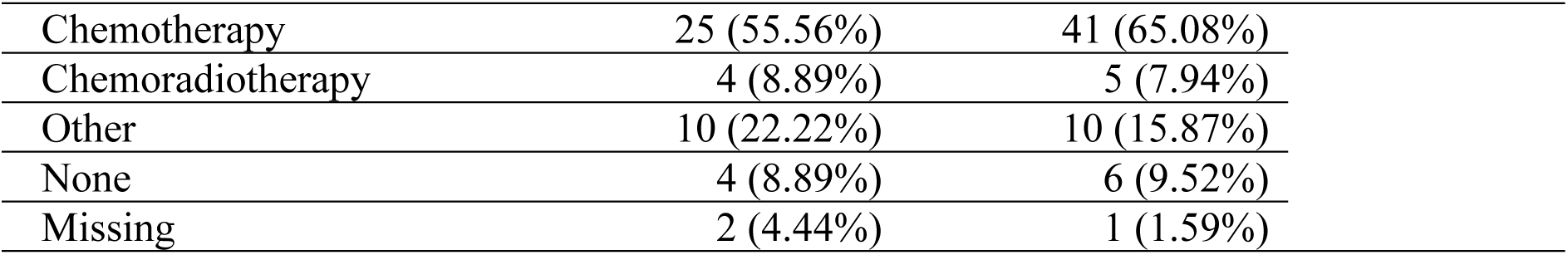
Characteristics of the study sample.

| **Variable** | **Improvers**<br>**(n=45)** | **No improvers**<br>**(n=63)** | **p-value** |
| --- | --- | --- | --- |
| **Age, mean (SD)** | 64.82 (9.59) | 63.91 (8.26) | 0.81 |
| **Sex, n (%)** |  |  |  |
| Male | 38 (84.44%) | 46 (73.02%) | 0.24 |
| Female | 7 (15.56%) | 17 (26.98%) |  |
| **BMI, mean (SD)** | 26.33 (5.00) | 26.94 (6.45) | 0.89 |
| **Smoking History, n (%)** |  |  |  |
| Ever smoked | 18 (40%) | 29 (46.03%) | 0.75 |
| Never smoked | 12 (26.67%) | 17 (26.98%) |  |
| Missing or Unknown | 15 (33.33%) | 17 (26.98%) |  |
| **Alcohol Use, n (%)** |  |  |  |
| Ever drank | 19 (42.22%) | 27 (42.86%) | 0.79 |
| Non-drinker | 10 (22.22%) | 17 (26.98%) |  |
| Missing or Unknown | 16 (35.56%) | 19 (30.16%) |  |
| **Charlson Comorbidity Index, mean (SD)** |  |  |  |
| none (0) | 0 (0%) | 1 (1.6%) | 0.79 |
| mild (1–2) | 7 (15.56%) | 12 (19.05%) |  |
| moderate (3–4) | 25 (55.56%) | 34 (53.97%) |  |
| severe (≥5) | 13 (28.89%) | 16 (25.40%) |  |
| **Surgery approach, n (%)** |  |  |  |
| Open | 25 | 36 | 0.71 |
| Minimally invasive | 18 | 22 |  |
| Missing | 2 | 5 |  |
| **Pathological stage, n (%)** |  |  |  |
| 0 | 1 (2.22%) | 5 (7.94%) | 0.59 |
| 1 | 2 (4.44%) | 1 (1.59%) |  |
| 2 | 6 (13.33%) | 10 (15.87%) |  |
| 3 | 8 (17.78%) | 10 (15.87%) |  |
| 4 | 3 (6.67%) | 2 (3.17%) |  |
| pCR | 0 (0%) | 2 (3.17%) |  |
| Missing or Unknown | 25 (55.56%) | 33 (52.38%) |  |
| **Tumor level, n (%)** |  |  |  |
| Cardia/GEJ | 22 (48.89%) | 28 (44.44%) | 0.40 |
| Lower Esophagus | 21 (46.67%) | 26 (41.27%) |  |
| Middle or Upper Esophagus | 2 (4.44%) | 8 (12.70%) |  |
| Missing or Unknown | 0 (0%) | 1 (1.59%) |  |
| **Histology, n (%)** |  |  |  |
| No Cancer | 5 (11.11%) | 7 (11.11%) | 0.22 |
| Adenocarcinoma | 30 (66.67%) | 33 (52.38%) |  |
| Squamous Cell Carcinoma | 1 (2.22%) | 9 (14.29%) |  |
| Other | 9 (20.00%) | 13 (20.63%) |  |
| Missing or Unknown | 0 (0%) | 1 (1.59%) |  |
| **Neoadjuvant treatment type, n (%)** |  |  | 0.78 |

|  |  |  |
| --- | --- | --- |
| Chemotherapy | 25 (55.56%) | 41 (65.08%) |
| Chemoradiotherapy | 4 (8.89%) | 5 (7.94%) |
| Other | 10 (22.22%) | 10 (15.87%) |
| None | 4 (8.89%) | 6 (9.52%) |
| Missing | 2 (4.44%) | 1 (1.59%) |

Differences in pre- and post-esophagectomy FACT-E scores between improvers and non-improvers are presented in Table 2. At baseline, patients who ultimately improved had significantly lower total FACT-E scores compared to non-improvers (mean = 116.43 vs. 137.68; p < 0.05), as well as significantly lower scores across all subdomains (PWB, SWB, EWB, FWB, ECS; all p < 0.05).

**Table 2.**
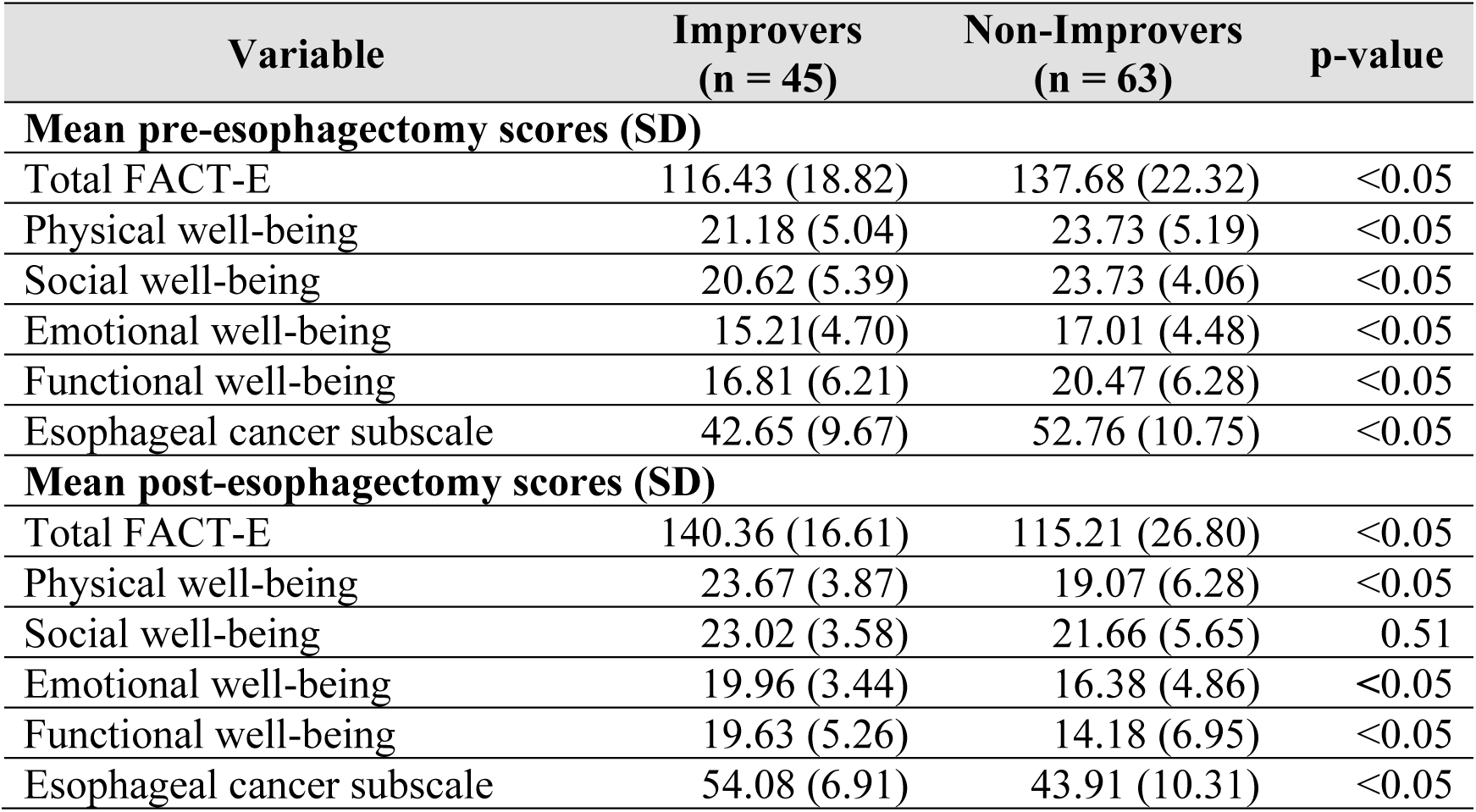
Pre- and post-esophagectomy FACT-E total and subdomain scores for improvers and non-improvers.

At one year post-esophagectomy, improvers demonstrated significantly higher total FACT-E scores than non-improvers (140.36 vs. 115.21; p < 0.05), along with higher scores in each subdomain except social well-being (SWB), which did not differ significantly between groups (p = 0.51). These results confirm that patients classified as “improvers” had both greater symptom burden at baseline and greater gains in HRQoL post-operatively.

### Univariate Associations Between Modifiable Factors and HRQoL Improvement

Results of the univariate logistic regression analyses of modifiable baseline factors and improvement in HRQoL are detailed in Table 3. Lower pre-esophagectomy total FACT-E scores were significantly associated with higher odds of improvement (OR = 0.95; 95% CI: 0.93–0.98; p < 0.05). All subdomain scores were inversely associated with HRQoL improvement, indicating that participants with greater baseline symptom burden were more likely to experience clinically meaningful gains.

**Table 3.** Results of the univariate models of quality-of-life improvement one-year post-esophagectomy.

| **Variable** | **Odds Ratio** | **95% CI** | **p-value** |
| --- | --- | --- | --- |
| **Pre-esophagectomy FACT-E** |  |  |  |
| *Total score* | 0.95 | 0.93 - 0.98 | <0.05 |
| *Physical well-being* | 0.90 | 0.83 - 0.98 | <0.05 |
| *Social well-being* | 0.87 | 0.80 - 0.95 | <0.05 |
| *Emotional well-being* | 0.92 | 0.84 - 1.00 | <0.05 |
| *Functional well-being* | 0.91 | 0.86 - 0.97 | <0.05 |
| *Esophageal cancer subscale* | 0.91 | 0.88 - 0.95 | <0.05 |
| **BMI** | 0.98 | 0.92 - 1.05 | 0.89 |
| **Smoking status** | 0.61 | 0.27 - 1.38 | 0.24 |
| **Alcohol use** | 0.94 | 0.43 - 2.03 | 0.87 |

BMI, smoking history, and alcohol use were not significantly associated with HRQoL improvement in univariate models (p > 0.05 for all).

### Multivariable Associations with One-Year HRQoL Improvement

Table 4 presents the results of the four stepwise multivariable logistic regression models examining factors associated with one-year HRQoL improvement. Across all models, lower pre-operative total FACT-E score remained a consistent and significant factor associated with improvement, indicating that participants with greater baseline symptom burden were more likely to experience meaningful post-esophagectomy gains.

**Table 4.** Results of the stepwise multivariable logistic regression models of quality-of-life improvement one-year post-esophagectomy.

| Variable | Model 1: Modifiable Only |  |  | Model 2: + Tumor |  |  | Model 3: + Treatment |  |  | Model 4: + Demographics |  |  |
| --- | --- | --- | --- | --- | --- | --- | --- | --- | --- | --- | --- | --- |
|  | Estimate | CI | p-value | Estimate | CI | p-value | Estimate | CI | p-value | Estimate | CI | p-value |
| **Pre-esophagectomy total FACT-E** | 0.95 | 0.93-0.97 | <0.05 | 0.95 | 0.93–0.98 | <0.05 | 0.95 | 0.92–0.97 | <0.05 | 0.94 | 0.91–0.97 | <0.05 |
| **BMI** | 1.00 | 0.92-1.07 | 0.89 | 0.99 | 0.91–1.08 | 0.86 | 1.00 | 0.91–1.09 | 0.91 | 0.98 | 0.88–1.08 | 0.62 |
| **Smoking status** | 0.42 | 0.16-1.08 | 0.07 | 0.38 | 0.14–1.03 | 0.06 | 0.46 | 0.15–1.36 | 0.16 | 0.32 | 0.10–1.05 | 0.06 |
| **Alcohol use** | 0.91 | 0.38-2.21 | 0.84 | 0.81 | 0.33–2.01 | 0.65 | 0.65 | 0.24–1.73 | 0.38 | 0.65 | 0.23–1.82 | 0.41 |
| **Tumour Level** |  |  |  |  |  |  |  |  |  |  |  |  |
| *Cardia/GEJ* |  |  |  | (Ref) | (Ref) | (Ref) | (Ref) | (Ref) | (Ref) | (Ref) | (Ref) | (Ref) |
| *Lower Esophagus* |  |  |  | 1.15 | 0.42–3.15 | 0.78 | 0.89 | 0.30-2.70 | 0.84 | 0.78 | 0.24–2.48 | 0.67 |
| *Middle or Upper Esophagus* |  |  |  | 0.76 | 0.09–6.17 | 0.79 | 0.36 | 0.04-3.71 | 0.39 | 0.25 | 0.02–3.64 | 0.31 |
| **Histology** |  |  |  |  |  |  |  |  |  |  |  |  |
| *No Cancer* |  |  |  | (Ref) | (Ref) | (Ref) | (Ref) | (Ref) | (Ref) | (Ref) | (Ref) | (Ref) |
| *Adenocarcinoma* |  |  |  | 2.28 | 0.53–9.80 | 0.27 | 3.72 | 0.72–19.30 | 0.12 | 3.50 | 0.62–19.64 | 0.16 |
| *Squamous Cell Carcinoma* |  |  |  | 0.78 | 0.05–11.71 | 0.86 | 0.45 | 0.02–11.61 | 0.63 | 0.33 | 0.01–9.76 | 0.52 |
| *Other* |  |  |  | 1.39 | 0.27–7.33 | 0.70 | 1.54 | 0.24–9.98 | 0.65 | 1.30 | 0.18–9.59 | 0.80 |
| **Surgery approach** |  |  |  |  |  |  |  |  |  |  |  |  |
| *Open* |  |  |  |  |  |  | (Ref) | (Ref) | (Ref) | (Ref) | (Ref) | (Ref) |
| *Minimally invasive* |  |  |  |  |  |  | 0.80 | 0.27–2.36 | 0.69 | 0.78 | 0.25–2.42 | 0.66 |
| **Neoadjuvant treatment type** |  |  |  |  |  |  |  |  |  |  |  |  |
| *None* |  |  |  |  |  |  | (Ref) | (Ref) | (Ref) | (Ref) | (Ref) | (Ref) |
| *Chemotherapy* |  |  |  |  |  |  | 0.43 | 0.07–2.88 | 0.39 | 0.45 | 0.07–2.92 | 0.40 |
| *Chemoradiotherapy* |  |  |  |  |  |  | 4.16 | 0.20–87.82 | 0.36 | 6.32 | 0.27–148.90 | 0.25 |
| *Other* |  |  |  |  |  |  | 2.55 | 0.34–19.35 | 0.37 | 3.98 | 0.48–33.03 | 0.20 |
| **Age** |  |  |  |  |  |  |  |  |  | Removed | Removed | Removed |
| **Sex** |  |  |  |  |  |  |  |  |  |  |  |  |
| *Male* |  |  |  |  |  |  |  |  |  | (Ref) | (Ref) | (Ref) |
| *Female* |  |  |  |  |  |  |  |  |  | 0.19 | 0.04–0.84 | <0.05 |
| **Charlson Comorbidity Index** |  |  |  |  |  |  |  |  |  |  |  |  |
| *None* |  |  |  |  |  |  |  |  |  | (Ref) | (Ref) | (Ref) |
| *Mild* |  |  |  |  |  |  |  |  |  | 1.89 | 0.49–7.27 | 0.35 |
| *Moderate* |  |  |  |  |  |  |  |  |  | Removed | Removed | Removed |
| *Severe* |  |  |  |  |  |  |  |  |  | 1.44 | 0.42–4.96 | 0.57 |

Note: Variables marked as “Removed” were excluded from the final model due to multicollinearity, defined as having a variance inflation factor (VIF) greater than 10.
Note: Pathological stage was excluded due to substantial missing data.

Smoking status approached statistical significance in Model 4, the fully adjusted model, (OR = 0.32; 95% CI: 0.10–1.05; p = 0.06), suggesting a possible trend toward reduced odds of improvement among ever-smokers. BMI and alcohol use were not significantly associated with HRQoL improvement in any model.

None of the tumor-related or treatment-related factors were significantly associated with one-year HRQoL improvement.

Among demographic and clinical characteristics, female sex was significantly associated with lower odds of improvement (OR = 0.19; 95% CI: 0.04–0.84; p = 0.03). Age and moderate comorbidity burden (i.e., CCI = 3–4) were excluded from the final model due to multicollinearity with other variables.

### Model Performance

Table 5 presents performance metrics for each multivariable logistic regression model. Discrimination, as measured by the AUC, improved with each stepwise addition of covariates from 0.79 in the modifiable-only model to 0.85 in the full model indicating better classification of patients who experienced HRQoL improvement. Pseudo R² also increased across models, from 0.19 to 0.31, suggesting progressive gains in model fit. Although the AIC was lowest in the modifiable-only model (129.38), the full model provided the best overall balance between discrimination and fit, supporting its use for interpreting associations with HRQoL improvement.

**Table 5.** Performance of the stepwise multivariable logistic regression models.

| **Model** | **AIC** | **AUC** | **Pseudo R<sup>2</sup>** |
| --- | --- | --- | --- |
| Model 1: Modifiable only | 129.38 | 0.79 | 0.19 |
| Model 2: + Tumor | 136.81 | 0.80 | 0.20 |
| Model 3: + Treatment | 135.95 | 0.83 | 0.26 |
| Model 4: Full model | 135.53 | 0.85 | 0.31 |

### Sensitivity Analyses

Sensitivity analyses using varying thresholds for HRQoL improvement (4–9 points) yielded consistent findings across models (Supplementary Tables 1–5). The direction and statistical significance of associations remained stable, indicating that the observed relationships were robust to changes in the threshold definition. No substantial differences were noted in the interpretation of results, reinforcing confidence in the overall conclusions.

## DISCUSSION

This study examined modifiable pre-operative factors associated with improvement in HRQoL one year after esophagectomy for esophageal cancer. Of the 108 participants, 42% experienced a clinically meaningful improvement in HRQoL one year after surgery over their pre-esophagectomy HRQoL. The results demonstrated that greater symptom burden prior to esophagectomy (i.e., lower total FACT-E score) was significantly associated with post-operative improvement. This was the only modifiable factor that was statistically significant. Model performance improved with the addition of tumor, treatment, and demographic variables.

The observations from this study are concordant with those by van der Wilk et al. (2023), who observed that patients with higher baseline HRQoL experienced more pronounced declines in outcomes over time following esophagectomy (21). While this raises the possibility of a ceiling effect, our analyses suggest a more nuanced relationship. Those participants in our study who improved started with significantly lower pre-operative HRQoL scores but achieved higher post-operative scores compared to non-improvers. This suggests not a ceiling effect, rather the role that pre-esophagectomy HRQoL plays in post-operative recovery. These results highlight the importance of ongoing HRQoL monitoring and support across all baseline levels.

Smoking status approached statistical significance as a variable associated with HRQoL improvement, with ever-smokers showing a trend toward lower odds of improvement. Although the association was not statistically significant, this finding is consistent with prior research linking smoking to worse surgical and functional outcomes in esophageal cancer. Several studies have shown that smoking is associated with increased post-operative complications, especially pulmonary issues, which may contribute to slower recovery and poorer physical functioning (22)(23)(24)(25). These complications may, in turn, affect patients’ ability to regain HRQoL after surgery. While further research is needed to clarify this relationship, our findings support continued emphasis on smoking cessation as part of pre-operative counselling and survivorship planning.

Alcohol consumption was not significantly associated with HRQoL improvement in our study. This finding appears inconsistent with existing literature, which has identified alcohol use, particularly heavy preoperative consumption, as a risk factor for increased postoperative complications and worse outcomes after esophagectomy and gastrointestinal surgeries more broadly (26) (27). One possible explanation is that our variable was binary (yes/no for any past or current use), which may not adequately capture the spectrum of alcohol exposure. Without distinguishing between heavy, moderate, light, or past use, meaningful dose-response relationships may be obscured. Future studies should aim to define alcohol use more precisely to clarify its potential role in postoperative HRQoL trajectories.

Preoperative BMI was not significantly associated with HRQoL improvement at one year. This finding is consistent with a previous population-based study in Sweden, which also reported no significant association between BMI and HRQoL six months post-esophagectomy (7). However, while BMI may not impact postoperative HRQoL, it has been associated with other important outcomes. Several studies suggest that extremes of BMI, particularly underweight and severely obese categories, are linked to increased rates of surgical complications and mortality (28). Yet, others have found no association between high BMI and adverse outcomes, and in some cases, even suggest protective effects for certain complications (29). Taken together, while our findings align with prior research regarding HRQoL, further investigation is needed to understand the broader implications of BMI across different postoperative outcomes.

Although these tumor- and treatment-related variables were not the primary focus of our analysis, they were included in the multivariable models to explore possible associations with HRQoL improvement. We did not observe significant associations between one-year HRQoL improvement and tumor level, histologic subtype, or surgical approach. Prior research has linked upper or middle tumor location, squamous histology, and open surgical approaches to poorer postoperative HRQoL; our differing findings may reflect variation in patient selection, follow-up intervals, or how outcomes were defined (7) (21) (30). In contrast, our findings regarding neoadjuvant therapy are consistent with previous literature, which has generally found no clear association between treatment modality and long-term HRQoL outcomes (31). Notably, sex emerged as a significant factor in our analysis, with females more likely to report no HRQoL improvement - an observation not consistently reported in earlier studies (7) (15). These results suggest that clinical variables may not be consistently associated with HRQoL recovery and highlight the need for further research to clarify their role.

Given the observational nature of this study, these results should be interpreted with caution. Several other limitations of this study should be considered. First, this was a single-center study with a relatively small sample size (n = 108), which may reduce the generalizability of our findings to broader esophagectomy populations. However, these real-world data are likely to reflect the distribution of patients seen in high volume esophageal cancer clinics. Second, selection bias is possible, as patients who did not initiate the FACT-E or had too many missing responses to generate valid scores at either timepoint were excluded. These excluded patients may have differed in clinically important ways, such as experiencing poorer outcomes, being less engaged in follow-up, or facing barriers to survey completion. This may lead to an overestimation of post-operative HRQoL in our sample. Third, survivorship bias is inherent in our analysis since we only included patients who survived to the 1-year follow-up. Patients who did not survive are not represented, which may skew our results toward better outcomes.

Despite its limitations, this study provides insight into potential factors associated with recovery. In particular, lower baseline HRQoL scores were significantly associated with greater odds of improvement, suggesting that patients with higher symptom burden may experience the most meaningful gains following surgery. These findings highlight an opportunity to use preoperative HRQoL as a tool for identifying patients who may benefit from enhanced support. In particular, patients with higher preoperative HRQoL may be at greater risk of decline and could require additional attention through targeted prehabilitation or psychosocial interventions to preserve their well-being after surgery.

## Data Availability

All data produced in the present work are contained in the manuscript

## Supplementary Figures

**Supplementary Table 1:**
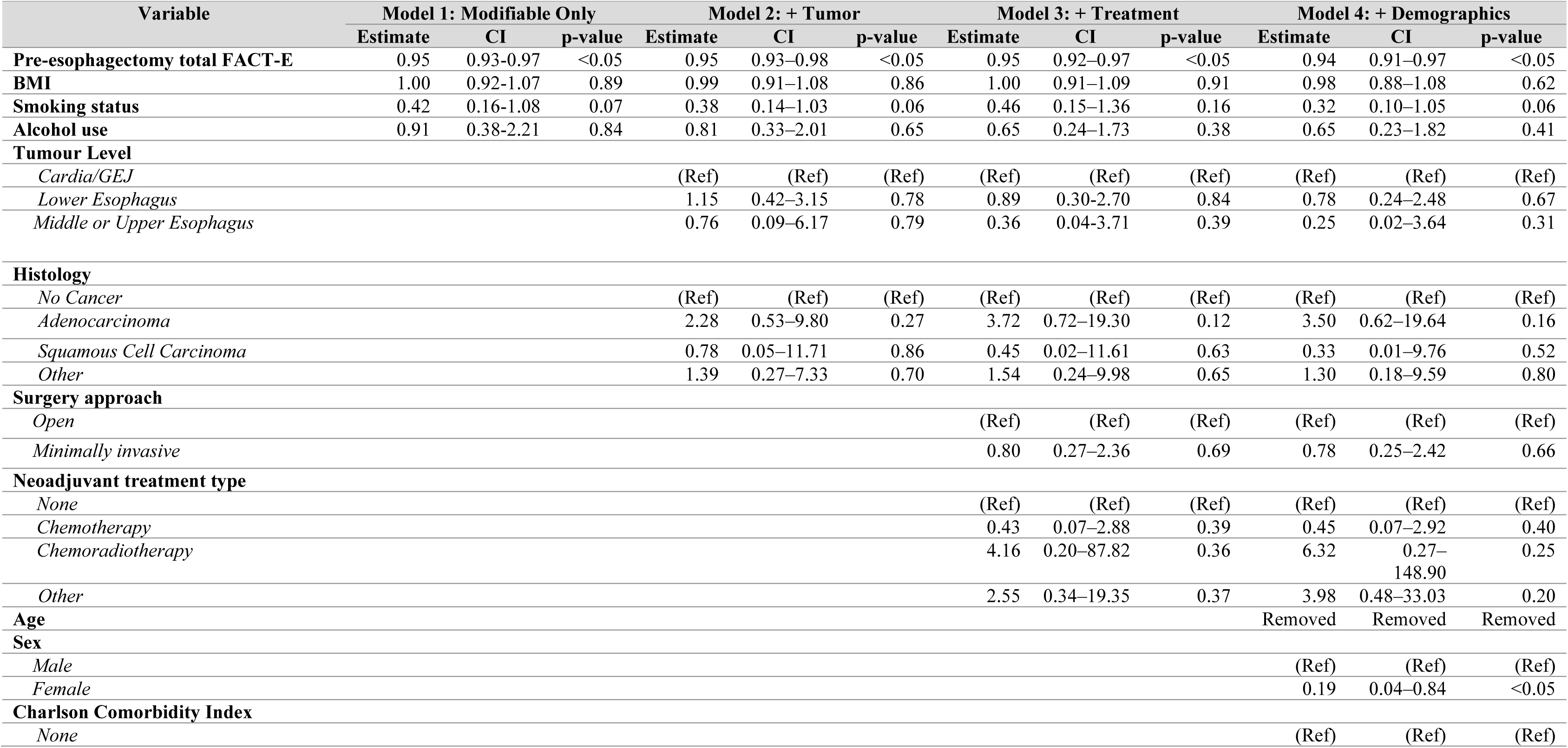
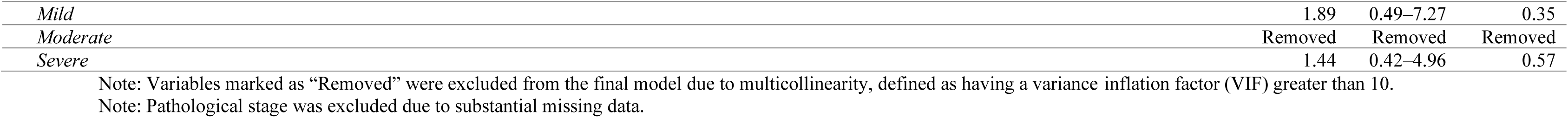
Results of the stepwise multivariable logistic regression models of quality-of-life improvement one-year post-esophagectomy (threshold = 4)

| Variable | Model 1: Modifiable Only |  |  | Model 2: + Tumor |  |  | Model 3: + Treatment |  |  | Model 4: + Demographics |  |  |
| --- | --- | --- | --- | --- | --- | --- | --- | --- | --- | --- | --- | --- |
|  | Estimate | CI | p-value | Estimate | CI | p-value | Estimate | CI | p-value | Estimate | CI | p-value |
| **Pre-esophagectomy total FACT-E** | 0.95 | 0.93-0.97 | <0.05 | 0.95 | 0.93–0.98 | <0.05 | 0.95 | 0.92–0.97 | <0.05 | 0.94 | 0.91–0.97 | <0.05 |
| **BMI** | 1.00 | 0.92-1.07 | 0.89 | 0.99 | 0.91–1.08 | 0.86 | 1.00 | 0.91–1.09 | 0.91 | 0.98 | 0.88–1.08 | 0.62 |
| **Smoking status** | 0.42 | 0.16-1.08 | 0.07 | 0.38 | 0.14–1.03 | 0.06 | 0.46 | 0.15–1.36 | 0.16 | 0.32 | 0.10–1.05 | 0.06 |
| **Alcohol use** | 0.91 | 0.38-2.21 | 0.84 | 0.81 | 0.33–2.01 | 0.65 | 0.65 | 0.24–1.73 | 0.38 | 0.65 | 0.23–1.82 | 0.41 |
| **Tumour Level** |  |  |  |  |  |  |  |  |  |  |  |  |
| *Cardia/GEJ* |  |  |  | (Ref) | (Ref) | (Ref) | (Ref) | (Ref) | (Ref) | (Ref) | (Ref) | (Ref) |
| *Lower Esophagus* |  |  |  | 1.15 | 0.42–3.15 | 0.78 | 0.89 | 0.30-2.70 | 0.84 | 0.78 | 0.24–2.48 | 0.67 |
| *Middle or Upper Esophagus* |  |  |  | 0.76 | 0.09–6.17 | 0.79 | 0.36 | 0.04-3.71 | 0.39 | 0.25 | 0.02–3.64 | 0.31 |
| **Histology** |  |  |  |  |  |  |  |  |  |  |  |  |
| *No Cancer* |  |  |  | (Ref) | (Ref) | (Ref) | (Ref) | (Ref) | (Ref) | (Ref) | (Ref) | (Ref) |
| *Adenocarcinoma* |  |  |  | 2.28 | 0.53–9.80 | 0.27 | 3.72 | 0.72–19.30 | 0.12 | 3.50 | 0.62–19.64 | 0.16 |
| *Squamous Cell Carcinoma* |  |  |  | 0.78 | 0.05–11.71 | 0.86 | 0.45 | 0.02–11.61 | 0.63 | 0.33 | 0.01–9.76 | 0.52 |
| *Other* |  |  |  | 1.39 | 0.27–7.33 | 0.70 | 1.54 | 0.24–9.98 | 0.65 | 1.30 | 0.18–9.59 | 0.80 |
| **Surgery approach** |  |  |  |  |  |  |  |  |  |  |  |  |
| *Open* |  |  |  |  |  |  | (Ref) | (Ref) | (Ref) | (Ref) | (Ref) | (Ref) |
| *Minimally invasive* |  |  |  |  |  |  | 0.80 | 0.27–2.36 | 0.69 | 0.78 | 0.25–2.42 | 0.66 |
| **Neoadjuvant treatment type** |  |  |  |  |  |  |  |  |  |  |  |  |
| *None* |  |  |  |  |  |  | (Ref) | (Ref) | (Ref) | (Ref) | (Ref) | (Ref) |
| *Chemotherapy* |  |  |  |  |  |  | 0.43 | 0.07–2.88 | 0.39 | 0.45 | 0.07–2.92 | 0.40 |
| *Chemoradiotherapy* |  |  |  |  |  |  | 4.16 | 0.20–87.82 | 0.36 | 6.32 | 0.27–148.90 | 0.25 |
| *Other* |  |  |  |  |  |  | 2.55 | 0.34–19.35 | 0.37 | 3.98 | 0.48–33.03 | 0.20 |
| **Age** |  |  |  |  |  |  |  |  |  | Removed | Removed | Removed |
| **Sex** |  |  |  |  |  |  |  |  |  |  |  |  |
| *Male* |  |  |  |  |  |  |  |  |  | (Ref) | (Ref) | (Ref) |
| *Female* |  |  |  |  |  |  |  |  |  | 0.19 | 0.04–0.84 | <0.05 |
| **Charlson Comorbidity Index** |  |  |  |  |  |  |  |  |  |  |  |  |
| *None* |  |  |  |  |  |  |  |  |  | (Ref) | (Ref) | (Ref) |

|  |  |  |  |
| --- | --- | --- | --- |
| *Mild* | 1.89 | 0.49–7.27 | 0.35 |
| *Moderate* | Removed | Removed | Removed |
| *Severe* | 1.44 | 0.42–4.96 | 0.57 |

Note: Variables marked as “Removed” were excluded from the final model due to multicollinearity, defined as having a variance inflation factor (VIF) greater than 10.
Note: Pathological stage was excluded due to substantial missing data.

**Supplementary Table 2:**
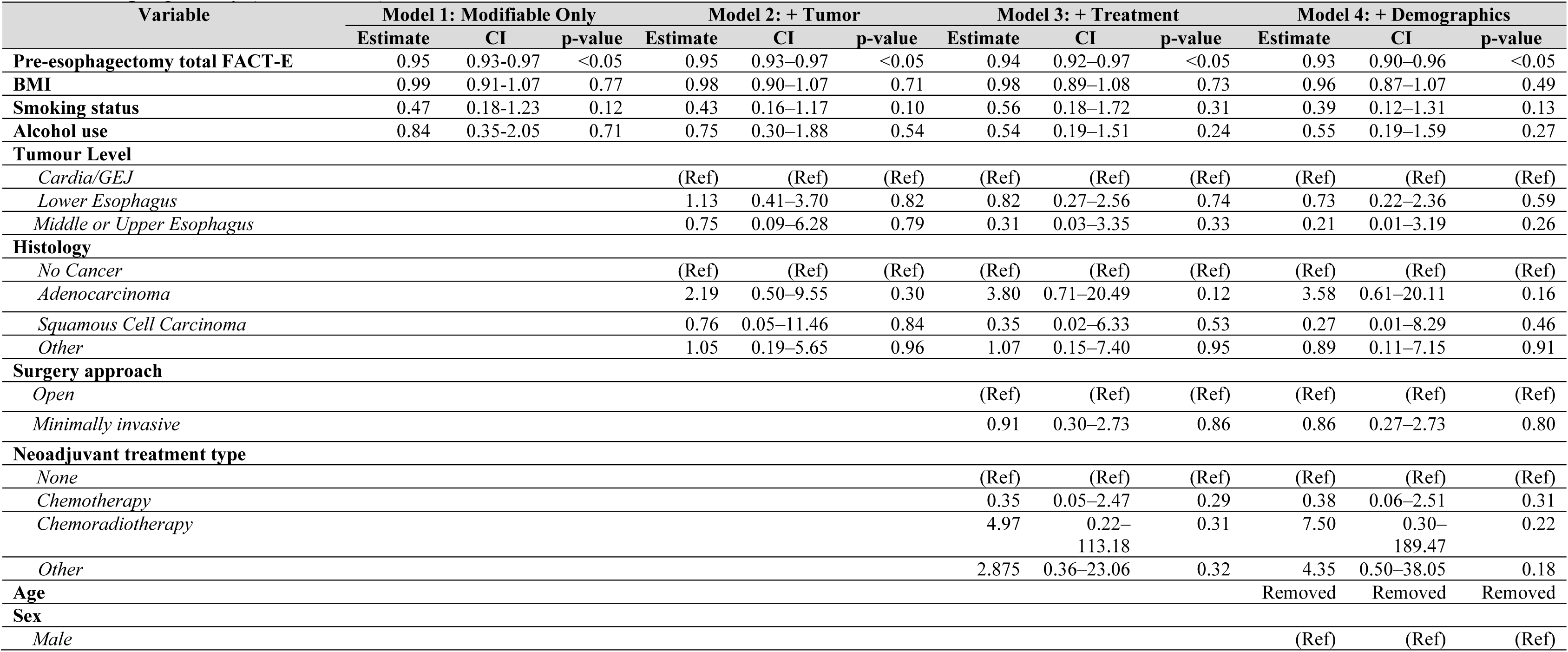
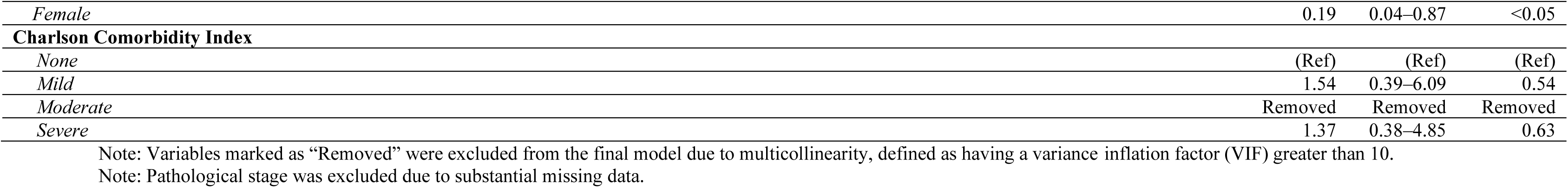
Results of the stepwise multivariable logistic regression models of quality-of-life improvement one-year post-esophagectomy (threshold = 6)

| Variable | Model 1: Modifiable Only |  |  | Model 2: + Tumor |  |  | Model 3: + Treatment |  |  | Model 4: + Demographics |  |  |
| --- | --- | --- | --- | --- | --- | --- | --- | --- | --- | --- | --- | --- |
|  | Estimate | CI | p-value | Estimate | CI | p-value | Estimate | CI | p-value | Estimate | CI | p-value |
| **Pre-esophagectomy total FACT-E** | 0.95 | 0.93-0.97 | <0.05 | 0.95 | 0.93–0.97 | <0.05 | 0.94 | 0.92–0.97 | <0.05 | 0.93 | 0.90–0.96 | <0.05 |
| **BMI** | 0.99 | 0.91-1.07 | 0.77 | 0.98 | 0.90–1.07 | 0.71 | 0.98 | 0.89–1.08 | 0.73 | 0.96 | 0.87–1.07 | 0.49 |
| **Smoking status** | 0.47 | 0.18-1.23 | 0.12 | 0.43 | 0.16–1.17 | 0.10 | 0.56 | 0.18–1.72 | 0.31 | 0.39 | 0.12–1.31 | 0.13 |
| **Alcohol use** | 0.84 | 0.35-2.05 | 0.71 | 0.75 | 0.30–1.88 | 0.54 | 0.54 | 0.19–1.51 | 0.24 | 0.55 | 0.19–1.59 | 0.27 |
| **Tumour Level** |  |  |  |  |  |  |  |  |  |  |  |  |
| *Cardia/GEJ* |  |  |  | (Ref) | (Ref) | (Ref) | (Ref) | (Ref) | (Ref) | (Ref) | (Ref) | (Ref) |
| *Lower Esophagus* |  |  |  | 1.13 | 0.41–3.70 | 0.82 | 0.82 | 0.27–2.56 | 0.74 | 0.73 | 0.22–2.36 | 0.59 |
| *Middle or Upper Esophagus* |  |  |  | 0.75 | 0.09–6.28 | 0.79 | 0.31 | 0.03–3.35 | 0.33 | 0.21 | 0.01–3.19 | 0.26 |
| **Histology** |  |  |  |  |  |  |  |  |  |  |  |  |
| *No Cancer* |  |  |  | (Ref) | (Ref) | (Ref) | (Ref) | (Ref) | (Ref) | (Ref) | (Ref) | (Ref) |
| *Adenocarcinoma* |  |  |  | 2.19 | 0.50–9.55 | 0.30 | 3.80 | 0.71–20.49 | 0.12 | 3.58 | 0.61–20.11 | 0.16 |
| *Squamous Cell Carcinoma* |  |  |  | 0.76 | 0.05–11.46 | 0.84 | 0.35 | 0.02–6.33 | 0.53 | 0.27 | 0.01–8.29 | 0.46 |
| *Other* |  |  |  | 1.05 | 0.19–5.65 | 0.96 | 1.07 | 0.15–7.40 | 0.95 | 0.89 | 0.11–7.15 | 0.91 |
| **Surgery approach** |  |  |  |  |  |  |  |  |  |  |  |  |
| *Open* |  |  |  |  |  |  | (Ref) | (Ref) | (Ref) | (Ref) | (Ref) | (Ref) |
| *Minimally invasive* |  |  |  |  |  |  | 0.91 | 0.30–2.73 | 0.86 | 0.86 | 0.27–2.73 | 0.80 |
| **Neoadjuvant treatment type** |  |  |  |  |  |  |  |  |  |  |  |  |
| *None* |  |  |  |  |  |  | (Ref) | (Ref) | (Ref) | (Ref) | (Ref) | (Ref) |
| *Chemotherapy* |  |  |  |  |  |  | 0.35 | 0.05–2.47 | 0.29 | 0.38 | 0.06–2.51 | 0.31 |
| *Chemoradiotherapy* |  |  |  |  |  |  | 4.97 | 0.22–<br>113.18 | 0.31 | 7.50 | 0.30–<br>189.47 | 0.22 |
| *Other* |  |  |  |  |  |  | 2.875 | 0.36–23.06 | 0.32 | 4.35 | 0.50–38.05 | 0.18 |
| **Age** |  |  |  |  |  |  |  |  |  | Removed | Removed | Removed |
| **Sex** |  |  |  |  |  |  |  |  |  |  |  |  |
| *Male* |  |  |  |  |  |  |  |  |  | (Ref) | (Ref) | (Ref) |

|  |  |  |  |
| --- | --- | --- | --- |
| *Female* | 0.19 | 0.04–0.87 | <0.05 |
| **Charlson Comorbidity Index** |  |  |  |
| *None* | (Ref) | (Ref) | (Ref) |
| *Mild* | 1.54 | 0.39–6.09 | 0.54 |
| *Moderate* | Removed | Removed | Removed |
| *Severe* | 1.37 | 0.38–4.85 | 0.63 |

Note: Variables marked as “Removed” were excluded from the final model due to multicollinearity, defined as having a variance inflation factor (VIF) greater than 10.
Note: Pathological stage was excluded due to substantial missing data.

**Supplementary Table 3:**
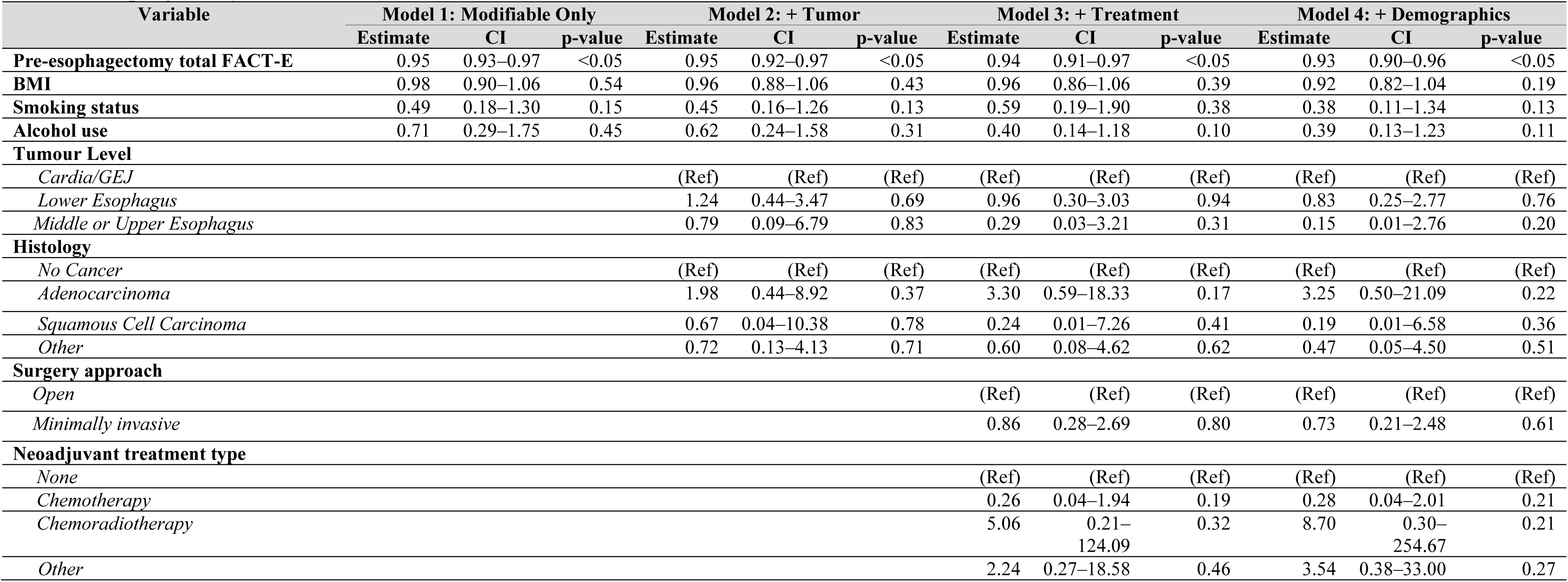
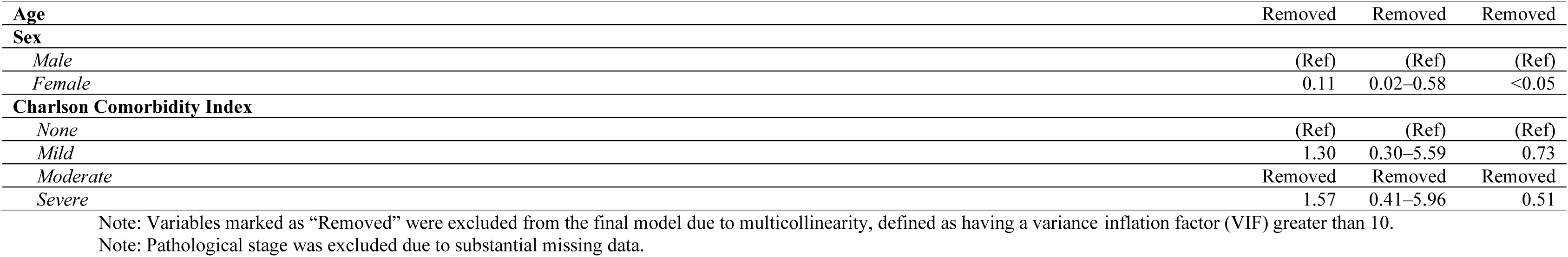
Results of the stepwise multivariable logistic regression models of quality-of-life improvement one-year post-esophagectomy (threshold = 7)

| Variable | Model 1: Modifiable Only |  |  | Model 2: + Tumor |  |  | Model 3: + Treatment |  |  | Model 4: + Demographics |  |  |
| --- | --- | --- | --- | --- | --- | --- | --- | --- | --- | --- | --- | --- |
|  | Estimate | CI | p-value | Estimate | CI | p-value | Estimate | CI | p-value | Estimate | CI | p-value |
| **Pre-esophagectomy total FACT-E** | 0.95 | 0.93–0.97 | <0.05 | 0.95 | 0.92–0.97 | <0.05 | 0.94 | 0.91–0.97 | <0.05 | 0.93 | 0.90–0.96 | <0.05 |
| **BMI** | 0.98 | 0.90–1.06 | 0.54 | 0.96 | 0.88–1.06 | 0.43 | 0.96 | 0.86–1.06 | 0.39 | 0.92 | 0.82–1.04 | 0.19 |
| **Smoking status** | 0.49 | 0.18–1.30 | 0.15 | 0.45 | 0.16–1.26 | 0.13 | 0.59 | 0.19–1.90 | 0.38 | 0.38 | 0.11–1.34 | 0.13 |
| **Alcohol use** | 0.71 | 0.29–1.75 | 0.45 | 0.62 | 0.24–1.58 | 0.31 | 0.40 | 0.14–1.18 | 0.10 | 0.39 | 0.13–1.23 | 0.11 |
| **Tumour Level** |  |  |  |  |  |  |  |  |  |  |  |  |
| *Cardia/GEJ* |  |  |  | (Ref) | (Ref) | (Ref) | (Ref) | (Ref) | (Ref) | (Ref) | (Ref) | (Ref) |
| *Lower Esophagus* |  |  |  | 1.24 | 0.44–3.47 | 0.69 | 0.96 | 0.30–3.03 | 0.94 | 0.83 | 0.25–2.77 | 0.76 |
| *Middle or Upper Esophagus* |  |  |  | 0.79 | 0.09–6.79 | 0.83 | 0.29 | 0.03–3.21 | 0.31 | 0.15 | 0.01–2.76 | 0.20 |
| **Histology** |  |  |  |  |  |  |  |  |  |  |  |  |
| *No Cancer* |  |  |  | (Ref) | (Ref) | (Ref) | (Ref) | (Ref) | (Ref) | (Ref) | (Ref) | (Ref) |
| *Adenocarcinoma* |  |  |  | 1.98 | 0.44–8.92 | 0.37 | 3.30 | 0.59–18.33 | 0.17 | 3.25 | 0.50–21.09 | 0.22 |
| *Squamous Cell Carcinoma* |  |  |  | 0.67 | 0.04–10.38 | 0.78 | 0.24 | 0.01–7.26 | 0.41 | 0.19 | 0.01–6.58 | 0.36 |
| *Other* |  |  |  | 0.72 | 0.13–4.13 | 0.71 | 0.60 | 0.08–4.62 | 0.62 | 0.47 | 0.05–4.50 | 0.51 |
| **Surgery approach** |  |  |  |  |  |  |  |  |  |  |  |  |
| *Open* |  |  |  |  |  |  | (Ref) | (Ref) | (Ref) | (Ref) | (Ref) | (Ref) |
| *Minimally invasive* |  |  |  |  |  |  | 0.86 | 0.28–2.69 | 0.80 | 0.73 | 0.21–2.48 | 0.61 |
| **Neoadjuvant treatment type** |  |  |  |  |  |  |  |  |  |  |  |  |
| *None* |  |  |  |  |  |  | (Ref) | (Ref) | (Ref) | (Ref) | (Ref) | (Ref) |
| *Chemotherapy* |  |  |  |  |  |  | 0.26 | 0.04–1.94 | 0.19 | 0.28 | 0.04–2.01 | 0.21 |
| *Chemoradiotherapy* |  |  |  |  |  |  | 5.06 | 0.21–124.09 | 0.32 | 8.70 | 0.30–254.67 | 0.21 |
| *Other* |  |  |  |  |  |  | 2.24 | 0.27–18.58 | 0.46 | 3.54 | 0.38–33.00 | 0.27 |

|  |  |  |  |
| --- | --- | --- | --- |
| **Age** | Removed | Removed | Removed |
| **Sex** |  |  |  |
| *Male* | (Ref) | (Ref) | (Ref) |
| *Female* | 0.11 | 0.02–0.58 | <0.05 |
| **Charlson Comorbidity Index** |  |  |  |
| *None* | (Ref) | (Ref) | (Ref) |
| *Mild* | 1.30 | 0.30–5.59 | 0.73 |
| *Moderate* | Removed | Removed | Removed |
| *Severe* | 1.57 | 0.41–5.96 | 0.51 |

Note: Variables marked as “Removed” were excluded from the final model due to multicollinearity, defined as having a variance inflation factor (VIF) greater than 10.
Note: Pathological stage was excluded due to substantial missing data.

**Supplementary Table 4:**
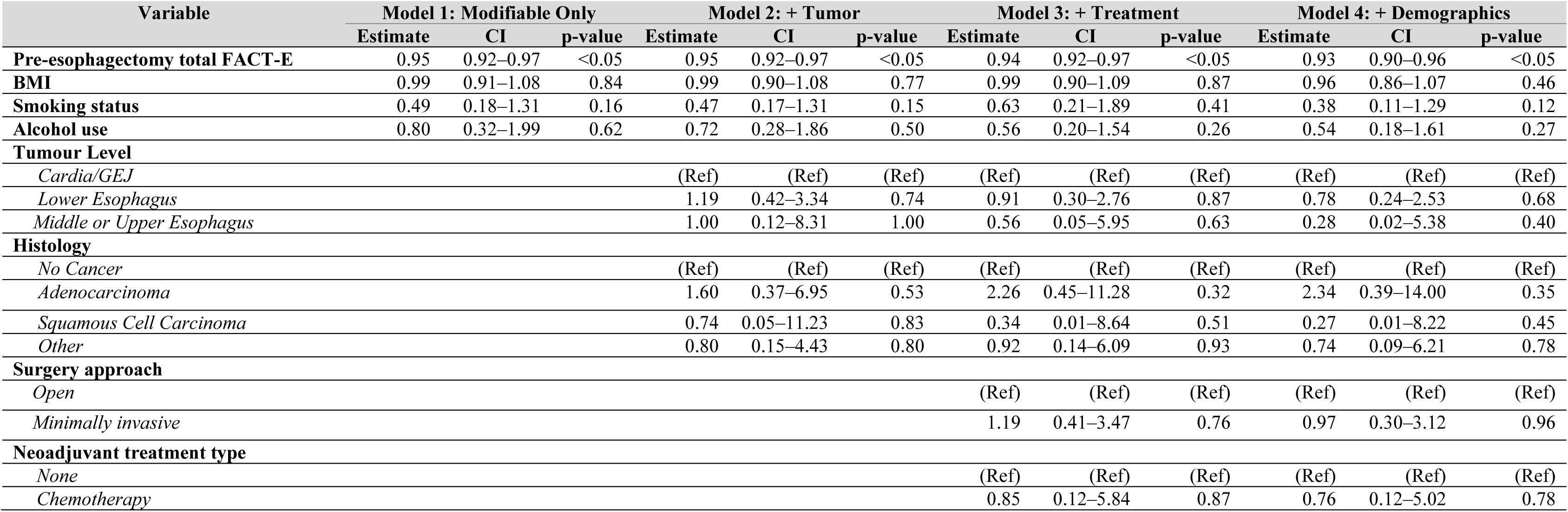
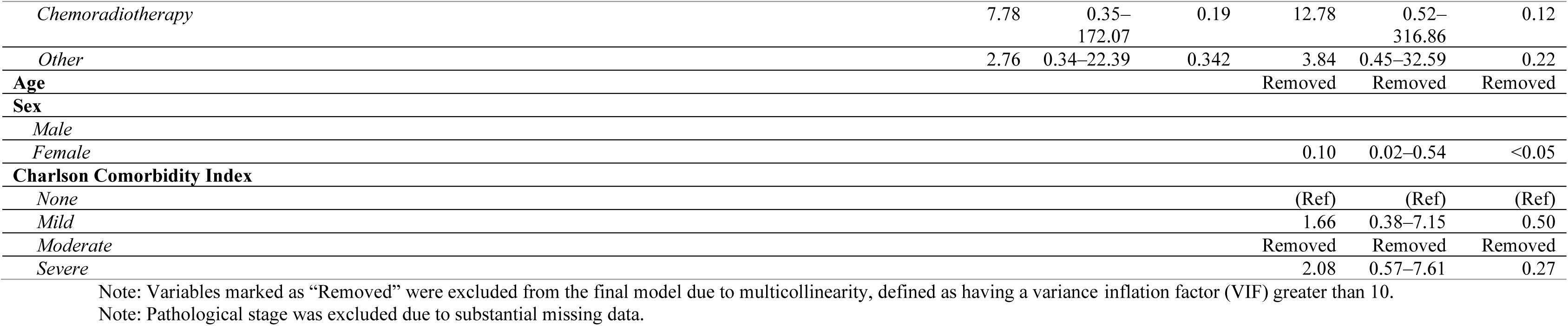
Results of the stepwise multivariable logistic regression models of quality-of-life improvement one-year post-esophagectomy (threshold = 8)

**Supplementary Table 5:**
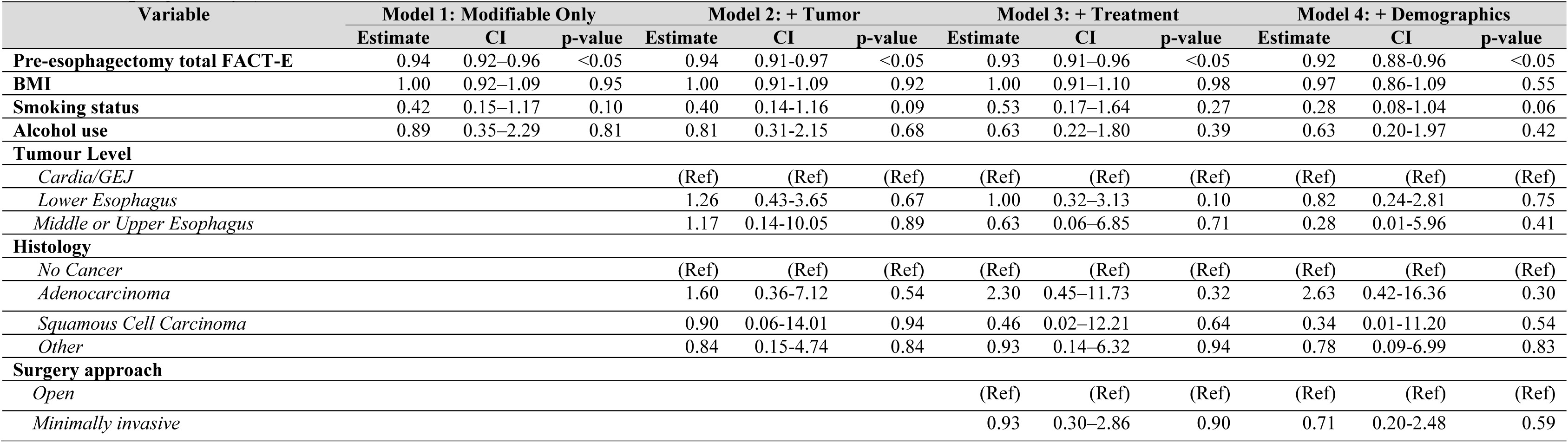
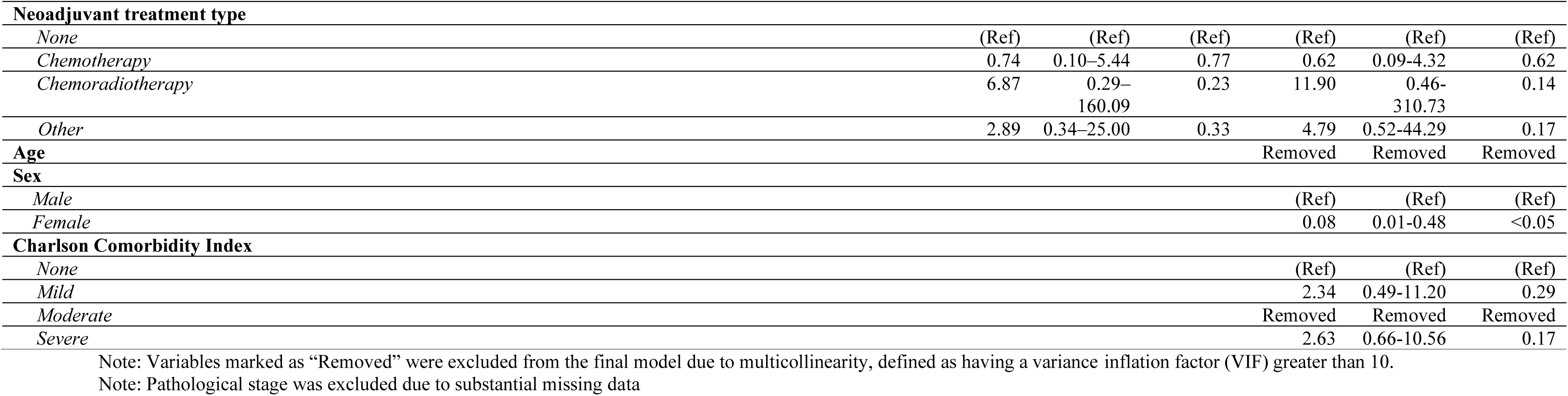
Results of the stepwise multivariable logistic regression models of quality-of-life improvement one-year post-esophagectomy (threshold = 9)

| Variable | Model 1: Modifiable Only |  |  | Model 2: + Tumor |  |  | Model 3: + Treatment |  |  | Model 4: + Demographics |  |  |
| --- | --- | --- | --- | --- | --- | --- | --- | --- | --- | --- | --- | --- |
|  | Estimate | CI | p-value | Estimate | CI | p-value | Estimate | CI | p-value | Estimate | CI | p-value |
| **Pre-esophagectomy total FACT-E** | 0.94 | 0.92–0.96 | <0.05 | 0.94 | 0.91-0.97 | <0.05 | 0.93 | 0.91–0.96 | <0.05 | 0.92 | 0.88-0.96 | <0.05 |
| **BMI** | 1.00 | 0.92–1.09 | 0.95 | 1.00 | 0.91-1.09 | 0.92 | 1.00 | 0.91–1.10 | 0.98 | 0.97 | 0.86-1.09 | 0.55 |
| **Smoking status** | 0.42 | 0.15–1.17 | 0.10 | 0.40 | 0.14-1.16 | 0.09 | 0.53 | 0.17–1.64 | 0.27 | 0.28 | 0.08-1.04 | 0.06 |
| **Alcohol use** | 0.89 | 0.35–2.29 | 0.81 | 0.81 | 0.31-2.15 | 0.68 | 0.63 | 0.22–1.80 | 0.39 | 0.63 | 0.20-1.97 | 0.42 |
| **Tumour Level** |  |  |  |  |  |  |  |  |  |  |  |  |
| *Cardia/GEJ* |  |  |  | (Ref) | (Ref) | (Ref) | (Ref) | (Ref) | (Ref) | (Ref) | (Ref) | (Ref) |
| *Lower Esophagus* |  |  |  | 1.26 | 0.43-3.65 | 0.67 | 1.00 | 0.32–3.13 | 0.10 | 0.82 | 0.24-2.81 | 0.75 |
| *Middle or Upper Esophagus* |  |  |  | 1.17 | 0.14-10.05 | 0.89 | 0.63 | 0.06–6.85 | 0.71 | 0.28 | 0.01-5.96 | 0.41 |
| **Histology** |  |  |  |  |  |  |  |  |  |  |  |  |
| *No Cancer* |  |  |  | (Ref) | (Ref) | (Ref) | (Ref) | (Ref) | (Ref) | (Ref) | (Ref) | (Ref) |
| *Adenocarcinoma* |  |  |  | 1.60 | 0.36-7.12 | 0.54 | 2.30 | 0.45–11.73 | 0.32 | 2.63 | 0.42-16.36 | 0.30 |
| *Squamous Cell Carcinoma* |  |  |  | 0.90 | 0.06-14.01 | 0.94 | 0.46 | 0.02–12.21 | 0.64 | 0.34 | 0.01-11.20 | 0.54 |
| *Other* |  |  |  | 0.84 | 0.15-4.74 | 0.84 | 0.93 | 0.14–6.32 | 0.94 | 0.78 | 0.09-6.99 | 0.83 |
| **Surgery approach** |  |  |  |  |  |  |  |  |  |  |  |  |
| *Open* |  |  |  |  |  |  | (Ref) | (Ref) | (Ref) | (Ref) | (Ref) | (Ref) |
| *Minimally invasive* |  |  |  |  |  |  | 0.93 | 0.30–2.86 | 0.90 | 0.71 | 0.20-2.48 | 0.59 |

|  |  |  |  |  |  |  |
| --- | --- | --- | --- | --- | --- | --- |
| Neoadjuvant treatment type |  |  |  |  |  |  |
| None | (Ref) | (Ref) | (Ref) | (Ref) | (Ref) | (Ref) |
| Chemotherapy | 0.74 | 0.10–5.44 | 0.77 | 0.62 | 0.09-4.32 | 0.62 |
| Chemoradiotherapy | 6.87 | 0.29–<br>160.09 | 0.23 | 11.90 | 0.46-310.73 | 0.14 |
| Other | 2.89 | 0.34–25.00 | 0.33 | 4.79 | 0.52-44.29 | 0.17 |
| Age |  |  |  | Removed | Removed | Removed |
| Sex |  |  |  |  |  |  |
| Male |  |  |  | (Ref) | (Ref) | (Ref) |
| Female |  |  |  | 0.08 | 0.01-0.48 | <0.05 |
| Charlson Comorbidity Index |  |  |  |  |  |  |
| None |  |  |  | (Ref) | (Ref) | (Ref) |
| Mild |  |  |  | 2.34 | 0.49-11.20 | 0.29 |
| Moderate |  |  |  | Removed | Removed | Removed |
| Severe |  |  |  | 2.63 | 0.66-10.56 | 0.17 |

Note: Variables marked as “Removed” were excluded from the final model due to multicollinearity, defined as having a variance inflation factor (VIF) greater than 10.
Note: Pathological stage was excluded due to substantial missing data

